# Weakly supervised learning for poorly differentiated adenocarcinoma classification in gastric endoscopic submucosal dissection whole slide images

**DOI:** 10.1101/2022.05.28.22275729

**Authors:** Masayuki Tsuneki, Fahdi Kanavati

## Abstract

The endoscopic submucosal dissection (ESD) is the preferred technique for treating early gastric cancers including poorly differentiated adenocarcinoma without ulcerative findings. The histopathological classification of poorly differentiated adenocarcinoma including signet ring cell carcinoma is of pivotal importance for determining further optimum cancer treatment(s) and clinical outcomes. Because conventional diagnosis by pathologists using microscopes is time-consuming and limited in terms of human resources, it is very important to develop computer-aided techniques that can rapidly and accurately inspect large numbers of histopathological specimen whole-slide images (WSIs). Computational pathology applications which can assist pathologists in detecting and classifying gastric poorly differentiated adenocarcinoma from ESD WSIs would be of great benefit for routine histopathological diagnostic workflow. In this study, we trained the deep learning model to classify poorly differentiated adenocarcinoma in ESD WSIs by transfer and weakly supervised learning approaches. We evaluated the model on ESD, endoscopic biopsy, and surgical specimen WSI test sets, achieving and ROC-AUC up to 0.975 in gastric ESD test sets for poorly differentiated adenocarcinoma. The deep learning model developed in this study demonstrates the high promising potential of deployment in a routine practical gastric ESD histopathological diagnostic workflow as a computer-aided diagnosis system.

## Introduction

According to the Global Cancer Statistics 2020, stomach cancer was responsible for 1,089,103 new cases (5.6% of all sites) and an estimated 768,793 deaths (7.7% of all sites) in 2020, ranking fifth for incidence and fourth for mortality globally^1^. So-called diffuse type adenocarcinoma (ADC) and signet ring cell carcinoma (SRCC) of stomach are poorly differentiated cancers (ADCs) which are believed to show poor prognosis and aggressive behavior^2^. Histopathologically, a diffuse growth of ADC cells is observed, associated with extensive fibrosis and inflammation and often the entire gastric wall is involved. Although a foveolar (intramucosal) type of SRCC occurs, in many cases of this entity the gastric mucosa is less affected than the deeper layers^3–5^. Therefore, poorly differentiated ADCs can often be mistaken for a variety of non-neoplastic lesions including gastritis, xanthoma/foamy histiocytes, or reactive endothelial cells in granulation tissues. Curative rates of endoscopic treatment for poorly differentiated type early gastric cancer are lower than those for the differentiated type ADC^6^. Endoscopic submucosal dissection (ESD) was developed in the late 1990s and has been widely used for early gastric cancer worldwide. ESD allows en bloc resection and precise histopathological inspection, while being a less invasive treatment than surgical resection^7^. Endoscopic resection is considered for tumors that have a very low possibility of lymph node metastasis and are suitable for en bloc resection^8^. According to the Japanese Gastric Cancer Treatment Guidelines 2018 (5th edition)^9^, the absolute indication of ESD is a differentiated type ADC with (UL1) or without (UL0) ulcerative findings and the expanded indication is a poorly differentiated (undifferentiated) ADC without ulcerative findings (UL0). Even in differentiated type ADC-dominant ESD specimens, it is important to inspect poorly differentiated ADC for the decision making of future therapeutic strategy^10^. Therefore, surgical pathologists are always on the lookout for signs of poorly differentiated ADC when evaluating gastric ESD.

In the field of computational pathology as a computer-aided detection (CADe) or computer-aided diagnosis (CADx), deep learning models have been widely applied in histopathological cancer classification on whole-slide images (WSIs), cancer cell detection and segmentation, and the stratification of patient clinical outcomes^11–24^. Previous studies have looked into applying deep learning models for ADC classification in stomach^24–26^, colon^24,27^, lung^25,28^, breast^29,30^, and prostate^31^ histopathological specimen WSIs. As for the gastric poorly differentiated ADC classification on WSIs, we have developed deep learning models based on endoscopic biopsy specimens^25,26^. However, the existing poorly differentiated ADC model did not classify poorly differentiated ADC precisely on gastric ESD WSIs.

In this study, we trained a deep learning model for the classification of gastric poorly differentiated ADC on ESD WSIs. We evaluated the trained model on ESD, endoscopic biopsy, and surgical specimen WSI test sets, achieving an ROC-AUC up to 0.975 in gastric ESD test sets, 0.960 in endoscopic biopsy test sets, and 0.929 in surgical specimen test sets. These findings suggest that deep learning models might be very useful as routine histopathological diagnostic aids for inspecting gastric ESD to detect poorly differentiated ADC precisely.

## Results

### High AUC performance of gastric ESD, biopsy, and surgical specimen WSI evaluation of gastric poorly differentiated ADC histopathology images

The aim of this retrospective study was to train a deep learning model for the classification of gastric poorly differentiated ADC in ESD WSIs. We have achieved high ROC-AUC performances in the ESD test sets (0.955 and 0.975) (Fig. 2A and Table 3). Prior to the training of gastric poorly differentiated ADC model using ESD WSIs (Table 1), we have demonstrated the existing gastric poorly differentiated ADC classification model (Biopsy-poorly ADC model)^25^ ROC-AUC performances on ESD, biopsy, and surgical specimen test sets (Table 2). As we have reported in the previous study^25^, the Biopsy-poorly ADC model achieved high ROC-AUC performances (0.959 - 0.976) in the biopsy test sets but not in the ESD and surgical test sets (Fig. 2, Table 3). However, to some extent, because the biopsy-poorly ADC model achieved moderately high ROC-AUC (0.899 and 0.638) in ESD test sets (Fig. 2, Table 3), we have trained the ESD-poorly ADC model based on the Biopsy-poorly ADC model using ESD training sets (Table 1).

**Table 1.** Distribution of gastric endoscopic submucosal dissection (ESD) whole-slide images (WSIs) in the training and validation sets obtained from the hospital-A.

| Hospital-A | Training sets | Validation sets | total |
| --- | --- | --- | --- |
| Poorly differentiated ADC | 140 | 10 | 150 |
| Differentiated ADC | 290 | 10 | 300 |
| Non-neoplastic lesion | 690 | 10 | 700 |
| total | 1120 | 30 | 1150 |

**Table 2.** Distribution of gastric endoscopic submucosal dissection (ESD), endoscopic biopsy, and surgical specimen whole-slide images in the test sets obtained from six hospitals (A-F)

| ESD-test set-1 (719 WSIs) |  |  |  |
| --- | --- | --- | --- |
| Supplier | Poorly differentiated ADC | Differentiated ADC | Non-neoplastic lesion |
| Hospital-A | 133 | 243 | 343 |
| ESD-test set-2 (637 WSIs) |  |  |  |
| Supplier | Poorly differentiated ADC | Differentiated ADC | Non-neoplastic lesion |
| Hospital-B | 7 | 69 | 54 |
| Hospital-C | 3 | 53 | 67 |
| Hospital-D | 11 | 89 | 78 |
| Hospital-E | 19 | 105 | 82 |
| Biopsy-test set-1 (355 WSIs) |  |  |  |
| Supplier | Poorly differentiated ADC | Differentiated ADC | Non-neoplastic lesion |
| Hospital-F | 25 | 96 | 234 |
| Biopsy-test set-2 (516 WSIs) |  |  |  |
| Supplier | Poorly differentiated ADC | Differentiated ADC | Non-neoplastic lesion |
| Hospital-B | 54 | 55 | 407 |
| Biopsy-test set-3 (495 WSIs) |  |  |  |
| Supplier | Poorly differentiated ADC | Differentiated ADC | Non-neoplastic lesion |
| Hospital-C | 10 | 12 | 473 |
| Biopsy-SRCC-test set (500 WSIs) |  |  |  |
| Supplier | SRCC | Differentiated ADC | Non-neoplastic lesion |
| Hospital-F | 54 | 55 | 391 |
| Surgical-test set (731 WSIs) |  |  |  |
| Supplier | Poorly differentiated ADC | Differentiated ADC | Non-neoplastic lesion |
| Hospital-B | 251 | 32 | 102 |
| Hospital-C | 211 | 24 | 111 |

**Table 3.** The comparison of ROC-AUC and log loss results for poorly differentiated adenocarcinoma (ADC) classification on various test sets between trained gastric endoscopic submucosal dissection (ESD) poorly differentiated ADC deep learning model (ESD-poorly ADC model) and existing biopsy model (Biopsy-poorly ADC model).

|  | ESD-poorly ADC model |  |
| --- | --- | --- |
|  | ROC-AUC | log-loss |
| ESD-test set-1 | 0.975 [0.962 - 0.986] | 0.235 [0.194 - 0.271] |
| ESD-test set-2 | 0.955 [0.915 - 0.981] | 0.658 [0.592 - 0.736] |
| Biopsy-test set-1 | 0.953 [0.909 - 0.981] | 0.352 [0.260 - 0.458] |
| Biopsy-test set-2 | 0.937 [0.892 - 0.969] | 0.205 [0.155 - 0.264] |
| Biopsy-test set-3 | 0.960 [0.922 - 0.989] | 0.156 [0.118 - 0.212] |
| Biopsy-SRCC-test set | 0.941 [0.896 - 0.975] | 0.185 [0.137 - 0.241] |
| Surgical-test set | 0.929 [0.909 - 0.949] | 0.572 [0.468 - 0.661] |

|  | Biopsy-poorly ADC model |  |
| --- | --- | --- |
|  | ROC-AUC | log-loss |
| ESD-test set-1 | 0.899 [0.869 - 0.927] | 0.857 [0.816 - 0.897] |
| ESD-test set-2 | 0.638 [0.541 - 0.722] | 1.104 [1.049 - 1.155] |
| Biopsy-test set-1 | 0.959 [0.931 - 0.982] | 0.587 [0.532 - 0.649] |
| Biopsy-test set-2 | 0.976 [0.951 - 0.993] | 0.199 [0.181 - 0.220] |
| Biopsy-test set-3 | 0.975 [0.946 - 0.994] | 0.418 [0.394 - 0.450] |
| Biopsy-SRCC-test set | 0.980 [0.953 - 0.995] | 0.190 [0.172 - 0.208] |
| Surgical-test set | 0.885 [0.857 - 0.910] | 0.518 [0.468 - 0.575] |

The models were applied in a sliding window fashion with an input tile size and stride of 224×224 pixels (Fig. 1). The transfer learning model (ESD-poorly ADC model) from existing Biopsy-poorly ADC model^25^ has higher ROC-AUCs, accuracy, sensitivitry, and specificity and lower log losses compared to the Biopsy-poorly ADC model in ESD and surgical specimen test sets but slightly lower ROC-AUCs compared to the Biopsy-poorly ADC model in biopsy test sets (Fig. 2, Tables 3, 4).

**Table 4.** The comparison of scores of accuracy, sensitivity, and specificity on the various test sets between trained gastric endoscopic submucosal dissection (ESD) poorly differentiated ADC deep learning model (ESD-poorly ADC model) and existing biopsy model (Biopsy-poorly ADC model).

|  | ESD-poorly ADC model |  |  |
| --- | --- | --- | --- |
|  | accuracy | sensitivity | specificity |
| ESD-test set-1 | 0.929 [0.912 - 0.949] | 0.910 [0.860 - 0.957] | 0.933 [0.915 - 0.954] |
| ESD-test set-2 | 0.940 [0.918 - 0.956] | 0.925 [0.829 - 1.000] | 0.941 [0.920 - 0.958] |
| Biopsy-test set-1 | 0.904 [0.870 - 0.935] | 0.920 [0.795 - 1.000] | 0.903 [0.869 - 0.936] |
| Biopsy-test set-2 | 0.882 [0.853 - 0.905] | 0.889 [0.786 - 0.962] | 0.881 [0.850 - 0.907] |
| Biopsy-test set-3 | 0.935 [0.911 - 0.954] | 0.900 [0.667 - 1.000] | 0.936 [0.911 - 0.955] |
| Biopsy-SRCC-test set | 0.884 [0.856 - 0.912] | 0.889 [0.786 - 0.964] | 0.883 [0.853 - 0.912] |
| Surgical-test set | 0.862 [0.837 - 0.889] | 0.875 [0.846 - 0.903] | 0.840 [0.795 - 0.884] |

|  | Biopsy-poorly ADC model |  |  |
| --- | --- | --- | --- |
|  | accuracy | sensitivity | specificity |
| ESD-test set-1 | 0.837 [0.811 - 0.865] | 0.790 [0.715 - 0.856] | 0.848 [0.820 - 0.877] |
| ESD-test set-2 | 0.776 [0.744 - 0.805] | 0.425 [0.256 - 0.567] | 0.799 [0.766 - 0.830] |
| Biopsy-test set-1 | 0.856 [0.820 - 0.890] | 0.960 [0.875 - 1.000] | 0.849 [0.809 - 0.885] |
| Biopsy-test set-2 | 0.921 [0.897 - 0.944] | 0.963 [0.904 - 1.000] | 0.916 [0.889 - 0.940] |
| Biopsy-test set-3 | 0.869 [0.838 - 0.897] | 0.900 [0.667 - 1.000] | 0.868 [0.838 - 0.897] |
| Biopsy-SRCC-test set | 0.936 [0.912 - 0.954] | 0.963 [0.895 - 1.000] | 0.933 [0.908 - 0.954] |
| Surgical-test set | 0.835 [0.804 - 0.859] | 0.855 [0.820 - 0.883] | 0.799 [0.747 - 0.845] |

**Figure 1.**
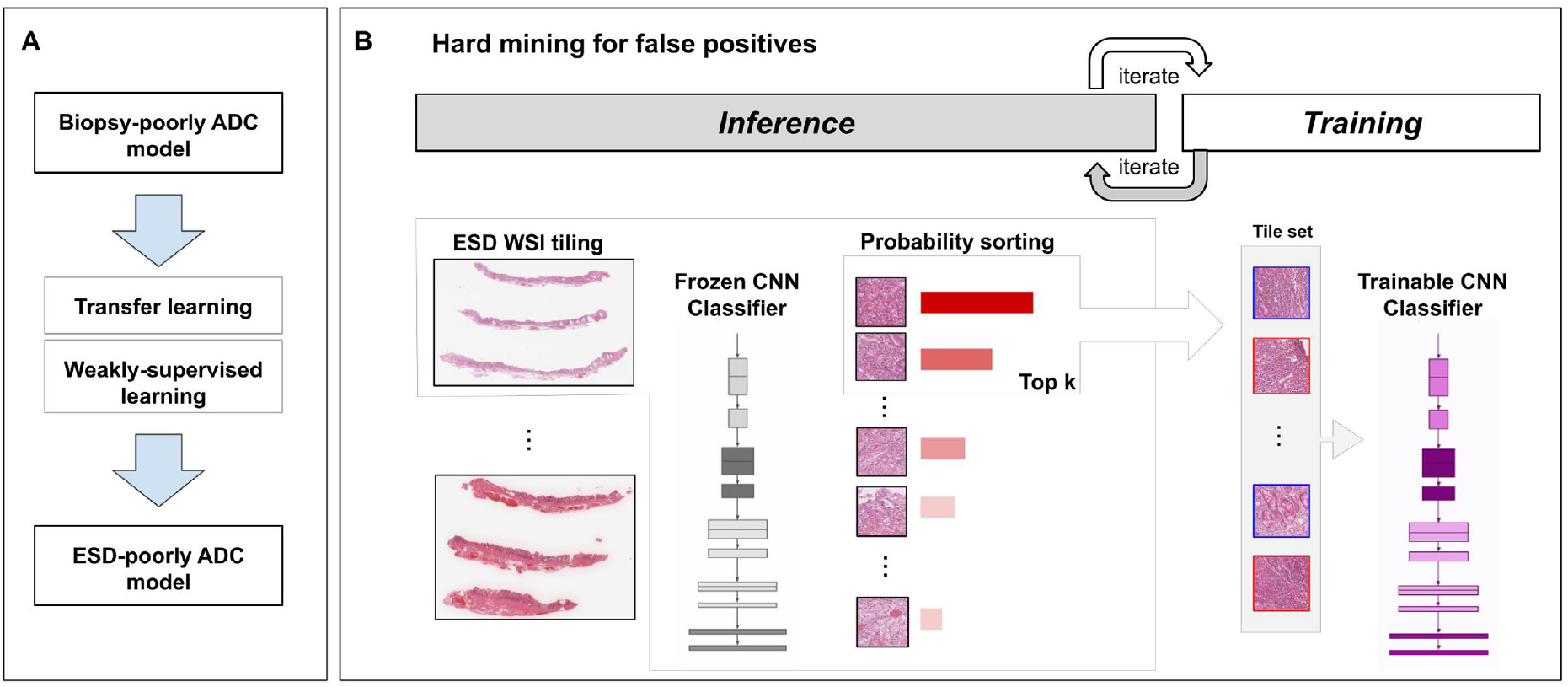
Schematic diagrams of training methods. (A) shows the simple summary of training method using transfer learning and weakly-supervised learning for this study. During training (B), we iteratively alternated between inference and training. During the inference step, the model weights were frozen and the model was used to select tiles with the highest probability after applying it on the entire tissue regions of each WSI. The top k tiles with the highest probabilities were then selected from each WSI and placed into a queue. During training, the selected tiles from multiple WSIs formed a training batch and were used to train the model.

**Figure 2.**
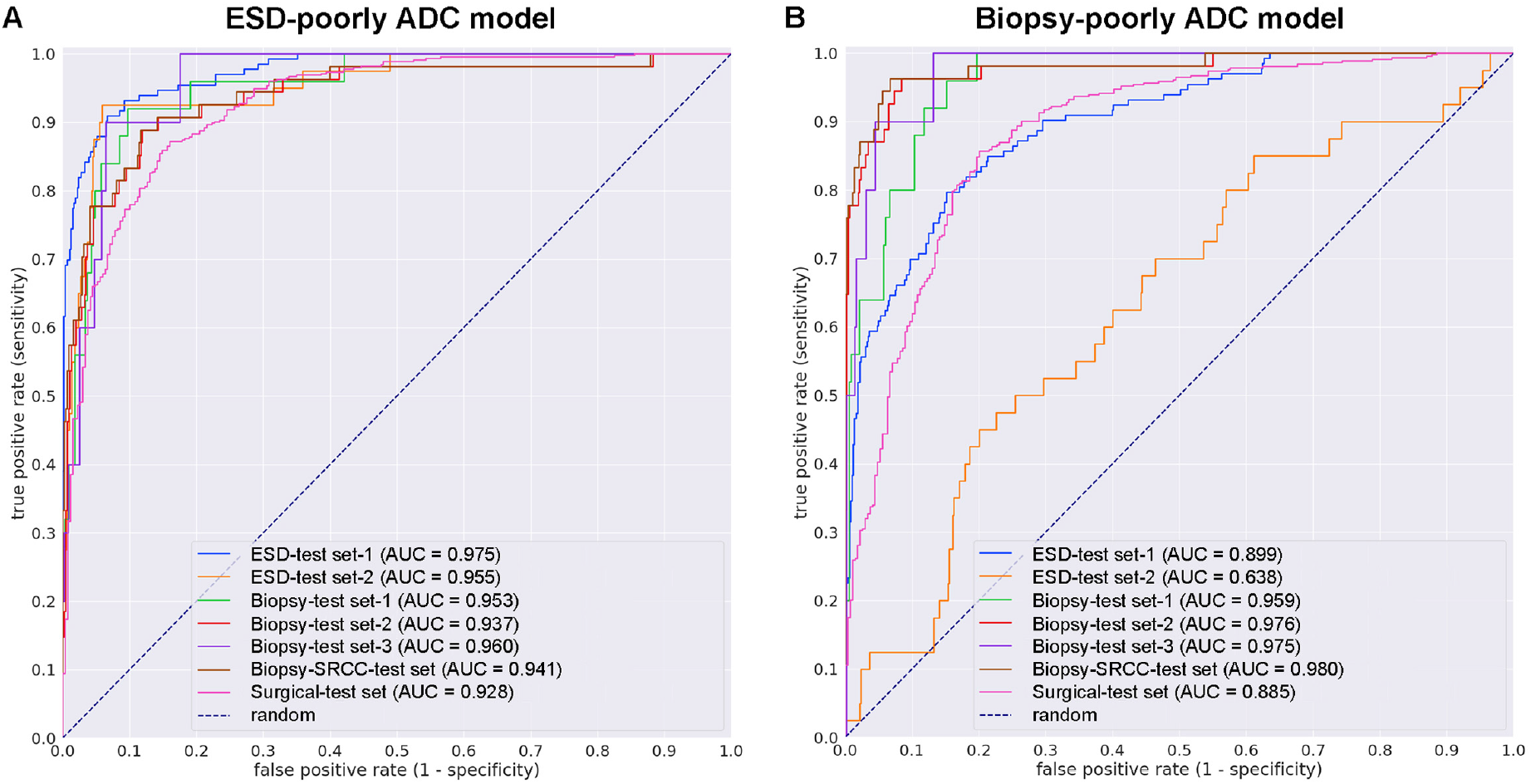
ROC curves with AUCs from trained gastric endoscopic submucosal dissection (ESD) poorly differentiated ADC deep learning model (ESD-poorly ADC model) and existing biopsy model (Biopsy-poorly ADC model) on the seven test sets: (A) the newly trained gastric ESD poorly differentiated ADC classification model (ESD-poorly ADC model) with tile size 224 px and magnification at x20; (B) the existing biopsy gastric poorly differentiated ADC classification model (Biopsy-poorly ADC model) with tile size 224 px and magnification at x20.

### True positive gastric poorly differentiated ADC prediction on ESD WSIs

Our model (ESD-poorly ADC model) satisfactorily predicted gastric poorly differentiated ADC in ESD WSIs (Fig. 3). According to the histopathological reports and additional pathologists’ reviewing, gastric poorly differentiated ADC cells were infiltrating in the neck area of gastric glands (Fig. 3A, C, E). The heatmap image (Fig. 3B) shows true positive predictions of poorly differentiated ADC cells (Fig. 3D, F) without false positive predictions in non-neoplastic areas. Histopathologically, there were trabecular and solid type poorly differentiated ADC cells which exhibited intramucosal invasive manners among differentiated type ADC with tubular or papillary structures (Fig. 3G, I). The heatmap image (Fig. 3H) shows true positive predictions of poorly differentiated ADC cells (Fig. 3J).

**Figure 3.**
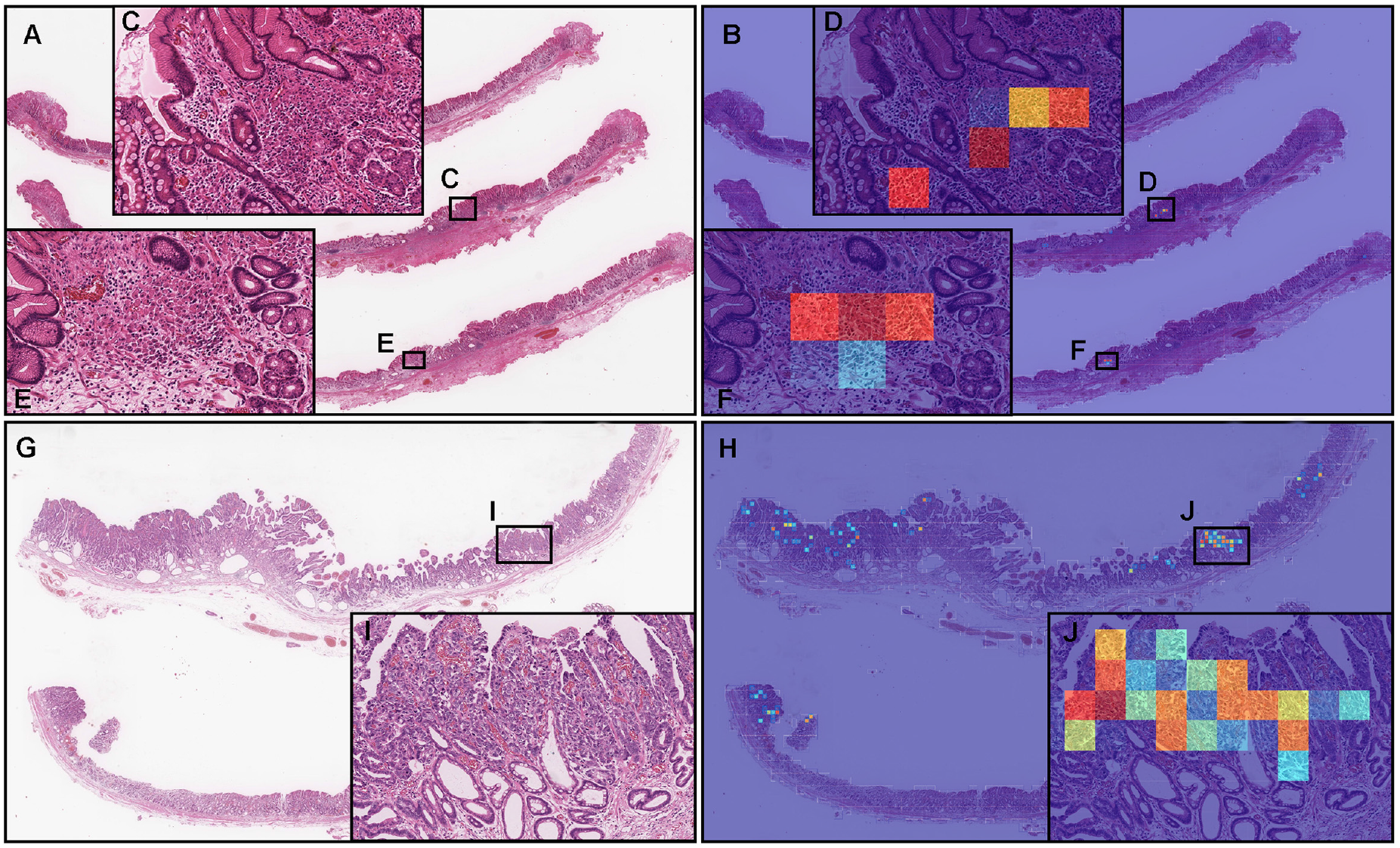
Two representative examples of poorly differentiated adenocarcinoma (ADC) true positive prediction outputs on whole-slide images (WSIs) from gastric endoscopic submucosal dissection (ESD) test sets using the model (ESD-poorly ADC model). In the gastric poorly differentiated ADC WSI of ESD specimen (A), poorly differentiated ADC cells were infiltrating in the neck area of gastric gland (C, E). The heatmap image (B) shows true positive predictions of gastric poorly differentiated ADC cells (D, F) which correspond respectively to H&E histopathology (C, E). The heatmap image (B) also shows no positive predictions (true negative predictions) in the tissue areas (A) without evidence of poorly differentiated ADC infiltration but with ulcerative gastritis. According to the histopathological report, (G) has differentiated and poorly differentiated ADC. In (I), there were gastric poorly differentiated ADC cells which exhibited trabecular and solid intramucosal invasive components. The heatmap image (H) shows true positive prediction of gastric poorly differentiated ADC cells (J) which correspond respectively to H&E histopathology (I). The heatmap uses the jet color map where blue indicates low probability and red indicates high probability.

### True negative gastric poorly differentiated ADC prediction on ESD WSIs

Our model (ESD-poorly ADC model) shows true negative predictions of gastric poorly differentiated ADC in ESD WSIs (Fig. 4A, B). Histopathologically, there was no evidence of presence of poorly differentiated ADC cells in all tissue fragments (#1-#3) which were non-neoplastic lesions with gastritis with ulcer formation (Fig. 4B) and were not predicted as gastric poorly differentiated ADC (Fig. 4C).

**Figure 4.**
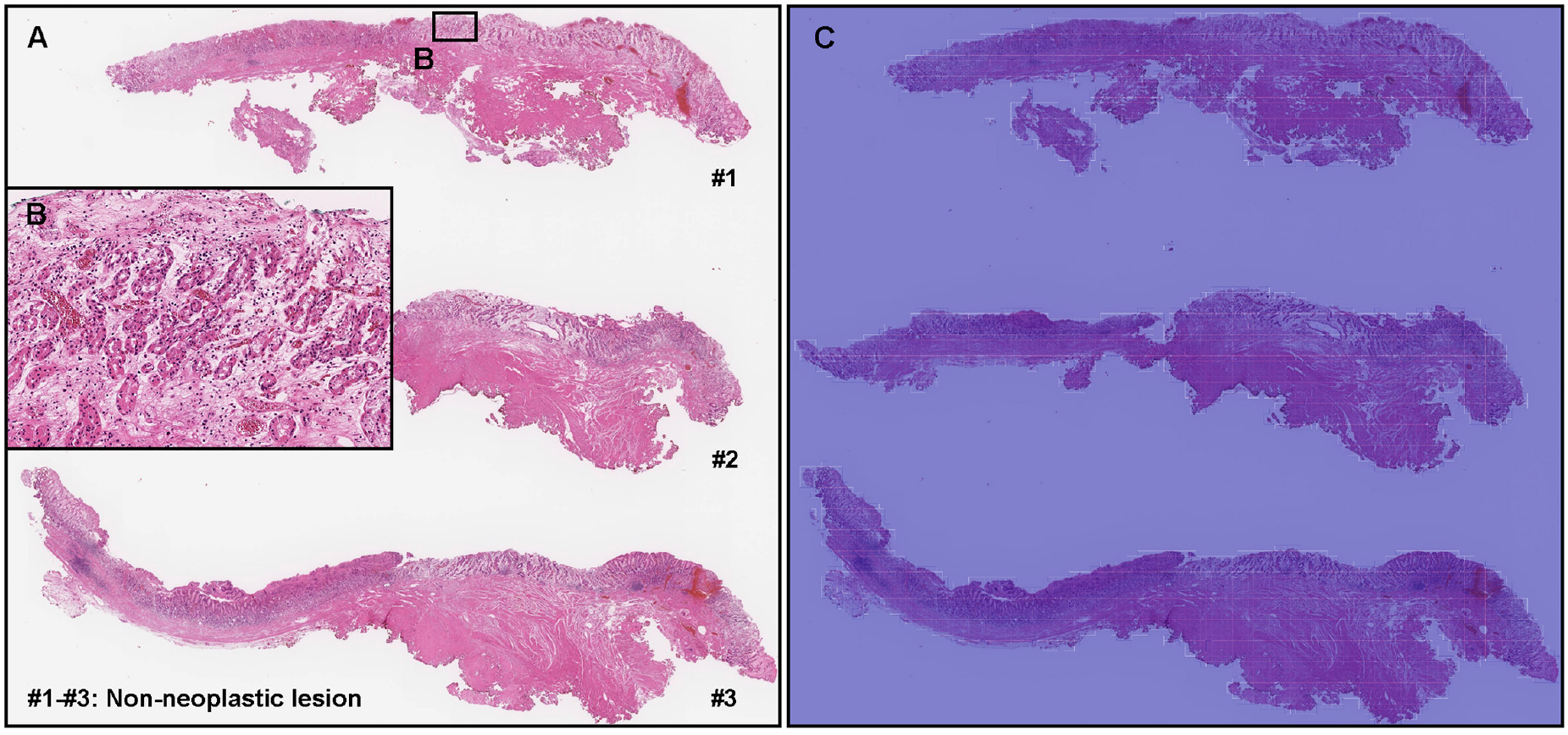
Representative true negative gastric poorly differentiated adenocarcinoma (ADC) prediction outputs on a whole slide image (WSI) from gastric endoscopic submucosal dissection (ESD) test sets using the model (ESD-poorly ADC model). Histopathologically, in (A), all tissue fragments (#1-#3) were non-neoplastic lesions with ulcerative gastritis (B). The heatmap image (C) shows true negative prediction of gastric poorly differentiated ADC. The heatmap uses the jet color map where blue indicates low probability and red indicates high probability.

### False positive gastric poorly differentiated ADC prediction on ESD WSIs

According to the histopathological report and additional pathologists’ reviewing, there were no gastric poorly differentiated ADC in these ESD fragments (#1-#3) which were non-neoplastic specimens (Fig. 5A). Our model (ESD-poorly ADC model) showed false positive predictions of poorly differentiated ADC (Fig. 5B). The false positively predicted areas (Fig. 5C, D) showed lymphatic tissue cells (e.g., lymphocyte, tingible body macrophage, and follicular dendritic cells) in artificially collapsed lymphoid follicle, which could be the primary cause of false positives due to its morphological similarity in poorly differentiated ADC cells.

**Figure 5.**
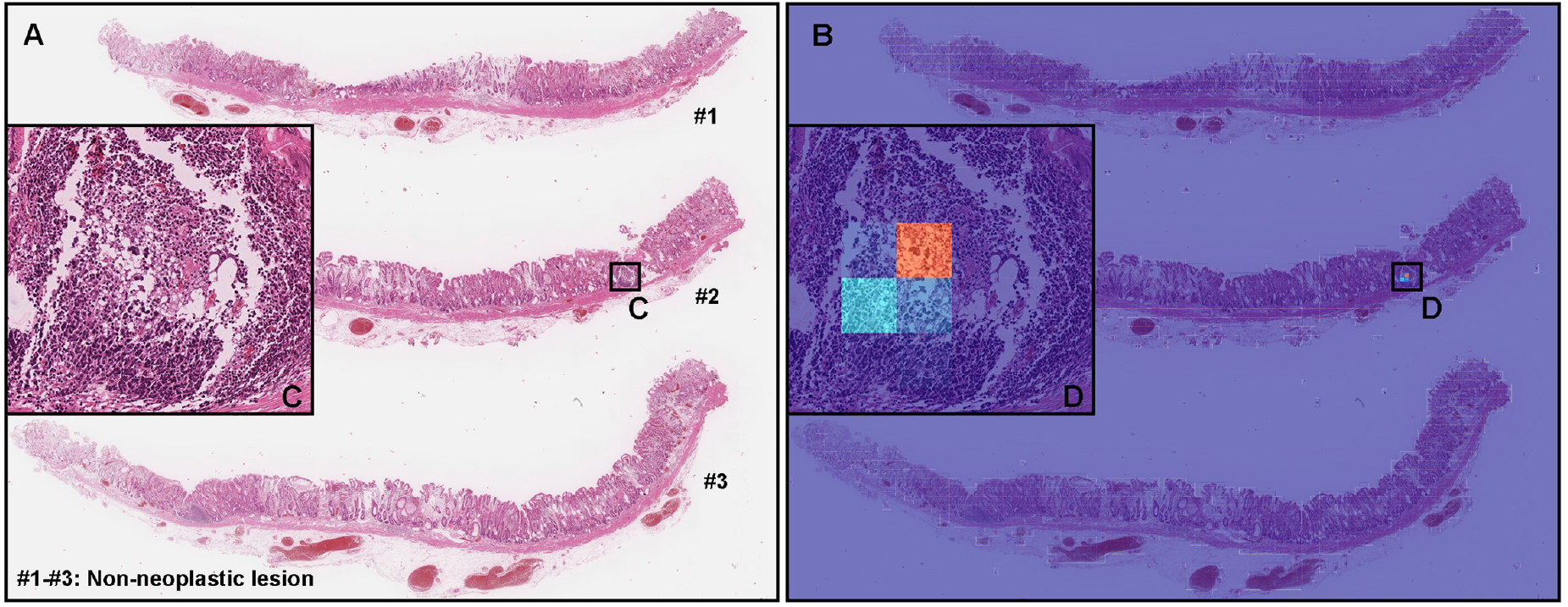
A representative example of gastric poorly differentiated adenocarcinoma (ADC) false positive prediction outputs on a whole slide image (WSI) from gastric endoscopic submucosal dissection (ESD) test sets using the model (ESD-poorly ADC model). Histopathologically, all tissue fragments (#1-#3) in (A) were non-neoplastic lesions. In the tissue fragment #2, The heatmap image (B) exhibits false positive predictions of gastric poorly differentiated ADC (D) on the lymphatic tissue cells in lymphoid follicle which was artificially collapsed during the specimen processing procedures. The heatmap uses the jet color map where blue indicates low probability and red indicates high probability.

### False negative gastric poorly differentiated ADC prediction on ESD WSIs

According to the histopathological report and additional pathologists’ reviewing, there were the foveolar-type signet ring cell carcinoma (SRCC) cells^3^ in the superficial layer of an ESD fragment (#1) (Fig. 6A, C) where pathologists marked with red-dots. Our model (ESD-poorly ADC model) did not predict poorly differentiated ADC cells (Fig. 6B, D). For comparison, we demonstrated predictions by our model (ESD-poorly ADC model) on 13 endoscopic biopsy WSIs with the presence of SRCC cells which have been false negatively predicted as SRCC by existing SRCC model^26^. Interestingly, there were 4 out of 13 WSIs with the presence of the foveolar-type SRCC cells which were also false negatively predicted as SRCC by our model (ESD-poorly ADC model). On the other hand, 9 out of 13 WSIs with the presence of SRCC cells were true positively predicted as poorly differentiated ADC by our model (ESD-poorly ADC model). Therefore, there is a limitation of our model (ESD-poorly ADC model) to predict the foveolar-type SRCC cells precisely.

**Figure 6.**
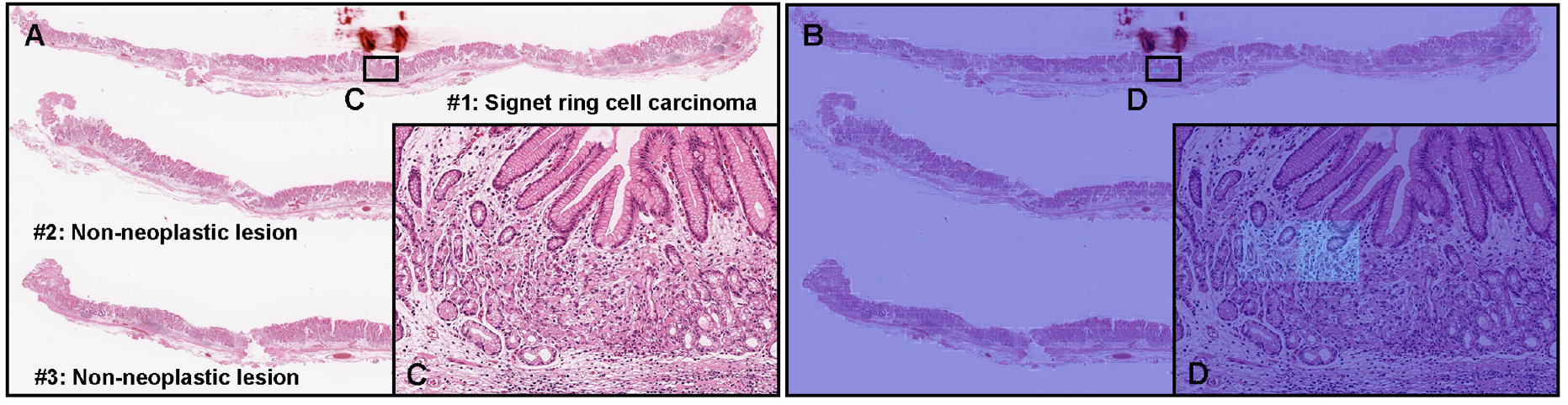
A representative example of gastric poorly differentiated adenocarcinoma (ADC) false negative prediction output on a whole slide image (WSI) from gastric endoscopic submucosal dissection (ESD) test sets using the model (ESD-poorly ADC model). Histopathologically, this case (A) has the foveolar-type signet ring cell carcinoma cell infiltration in the small area of superficial layer in the fragment #1 (C). The other two tissue fragments (#2, #3) were non-neoplastic lesions (A). The heatmap image (B) exhibited no positive poorly differentiated ADC prediction (D). The heatmap uses the jet color map where blue indicates low probability and red indicates high probability.

### True positive and true negative gastric poorly differentiated ADC prediction on surgical specimen WSIs

In addition, we have applied the model (ESD-poorly ADC model) on the surgical specimen WSIs. Figure 7 shows an example of surgical gastric poorly differentiated ADC case with five serial section WSIs (#1-#5) (Fig. 7A, C, E, G, I). We see the model (ESD-poorly ADC model) was capable of true positive (#1, #2, #4, #5) (Fig. 7A-D, G-J) and true negative (#3) (Fig. 7E, F) poorly differentiated ADC detection on such section WSIs. Histopathologically, gastric poorly differentiated ADC cells invading areas (Fig. 7K, M, O, Q) were visualized by heatmap images (Fig. 7L, N, P, R).

**Figure 7.**
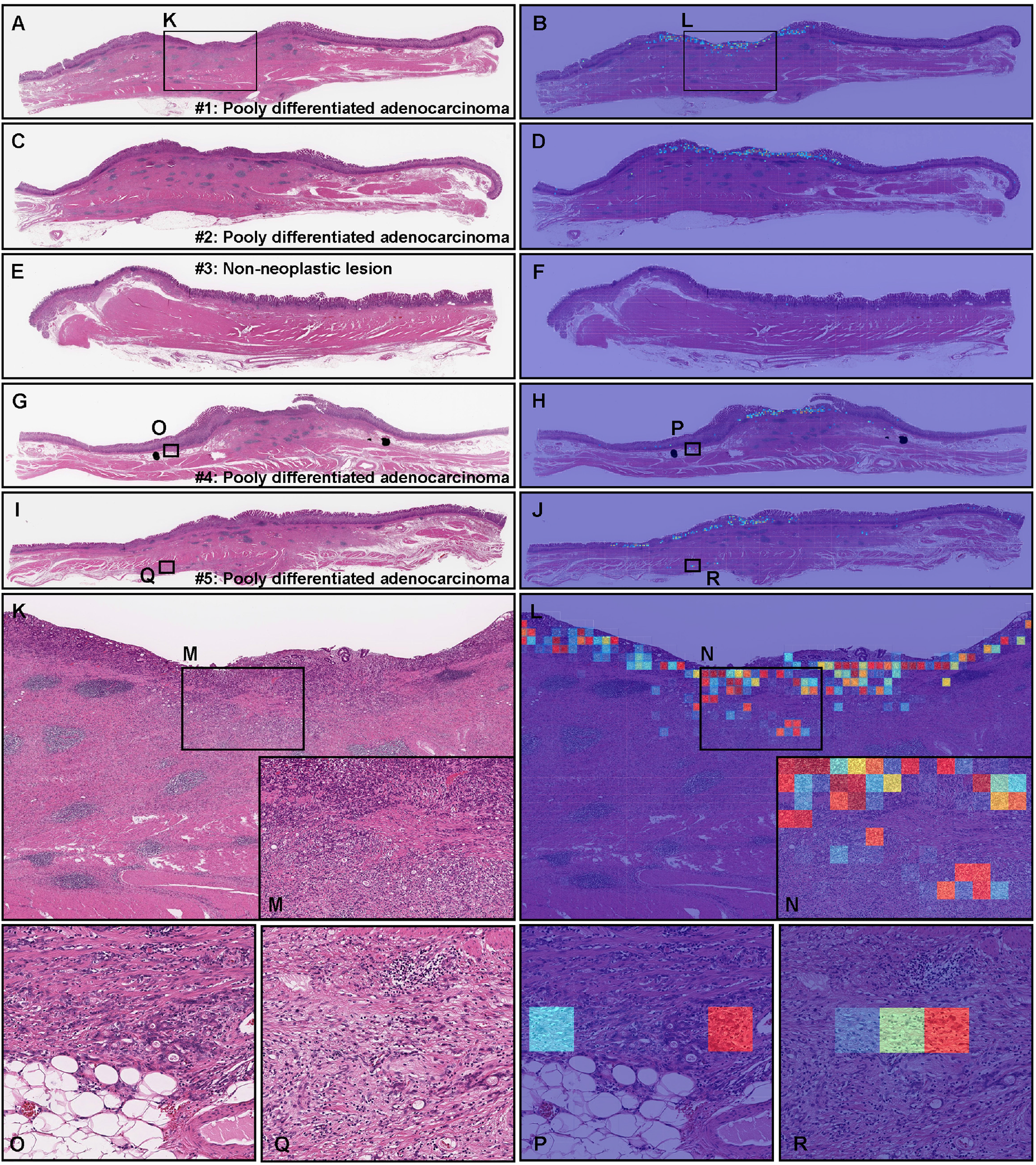
A representative surgically resected gastric poorly differentiated adenocarcinoma (ADC) case serial specimens. According to the histopathological report, serial specimen #1 (A), #2 (C), #4 (G), and #5 (I) have poorly differentiated ADC and #3 (E) is non-neoplastic lesion. The heatmap images show true positive predictions of gastric poorly differentiated ADC cells (B, D, H, J, L, N, P, and R) which correspond respectively to H&E histopathology (A, C, G, I, K, M, O, and Q) and true negative prediction (F). The heatmap uses the jet color map where blue indicates low probability and red indicates high probability.

### The application of deep learning models to classify gastric poorly differentiated ADC on various type of specimen

Based on the findings in this study and previous study^25^, we have summarized the possible application of our deep learning models (ESD-poorly ADC model and Biopsy-poorly ADC model) for classification of gastric poorly differentiated ADC in various type of specimen WSIs (Fig. 8). We can apply the model (ESD-poorly ADC model) for all types of specimen (ESD, biopsy, and surgical specimens), however, for biopsy specimen, the biopsy model (Biopsy-poorly ADC model) achieved slightly better performance than the ESD model (ESD-poorly ADC model).

**Figure 8.**
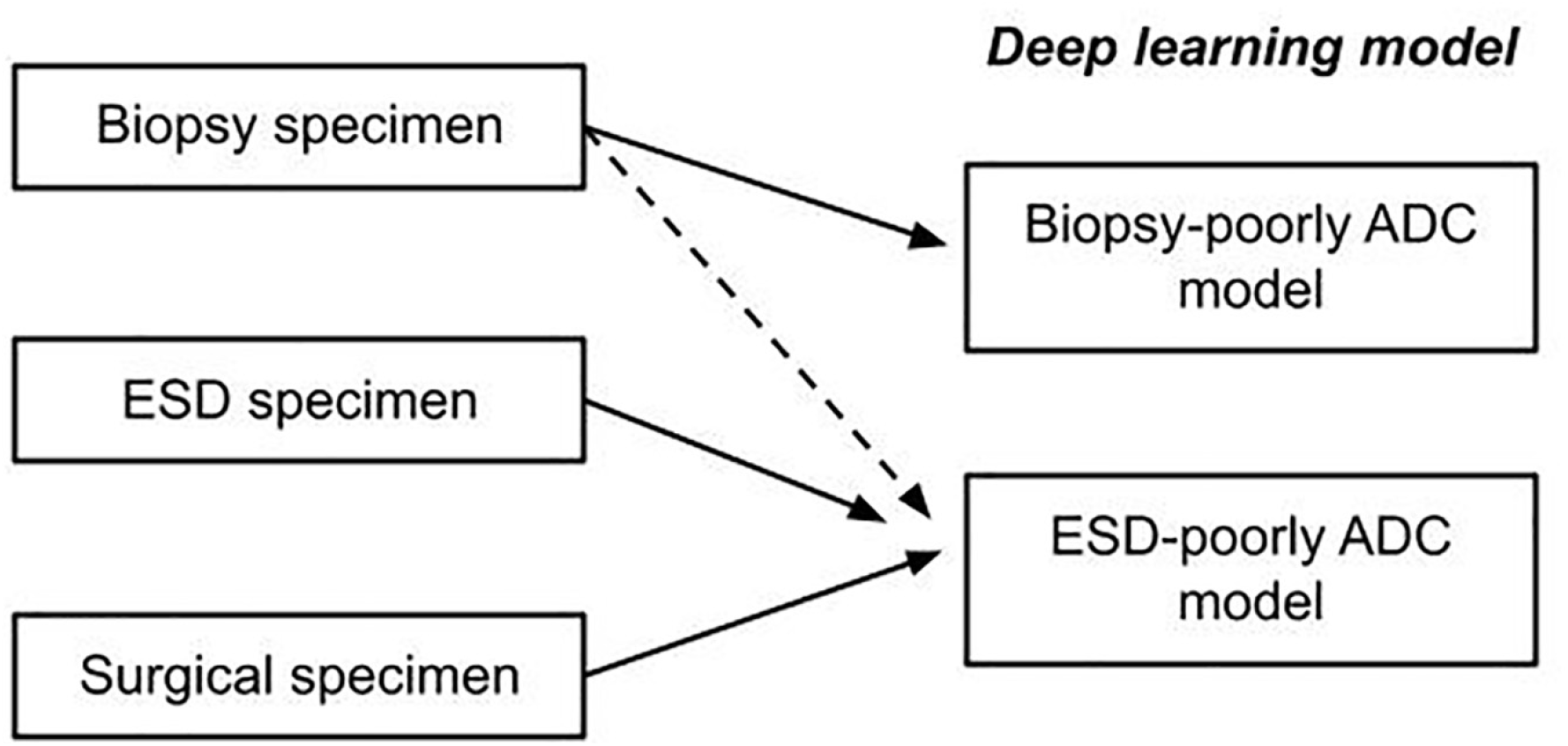
The schematic diagram of possible application of deep learning models (ESD-poorly ADC model and Biopsy-poorly ADC model) for classification of gastric poorly differentiated adenocarcinoma (ADC) in different type of specimens. ESD-poorly ADC model can be applied to classify gastric poorly differentiated ADC in biopsy, endoscopic submucosal dissection (ESD), and surgical specimen. Biopsy-poorly ADC model can be applied only in biopsy specimen. To classify gastric poorly differentiated ADC in biopsy specimen, both ESD-poorly ADC model and Biopsy-poorly ADC model can be used, however, Biopsy-poorly ADC model would be able to achieve higher ROC-AUC performance than ESD-poorly ADC model.

## Discussion

In this study, we trained a deep learning model for the classification of gastric poorly differentiated ADC in gastric ESD WSIs. Indications for gastric ESD were determined by presence or absence of a risk of nodal metastasis and according to the gastric cancer treatment guidelines^9,10^. As an expanded indication, the gastric poorly differentiated ADC without ulcerative findings (UL0) in which the depth of invasion is clinically diagnosed as T1a (cT1a) and the diameter is ≤2 cm can be endoscopically resected^9,32^. However, the mixed type tumors (poorly differentiated adenocarcinoma and SRCC) should not be considered for endoscopic resection due to the higher risk of nodal metastases^33^. According to the guidelines for ESD in early gastric cancer treatment by Japan Gastroenterological Endoscopy Society (JGES), differentiated and poorly differentiated mixed type ADCs measuring ≤2 cm in diameter with UL0 and cT1a are absolute indications for ESD^34^. Importantly, incidental gastric poorly differentiated ADCs are diagnosed at the time of ESD for differentiated type ADC, even though histopathological type was determined by endoscopic biopsy prior to ESD^10^. The histopathological evaluation of gastric ESD specimens whether there are poorly differentiated ADC cells or not is important for future therapeutic strategy because of lower rate of adverse events and high rate of en bloc resection^10^. Prior to training the deep learning model for ESD WSIs, we evaluated the ROC-AUC on gastric ESD test sets using existing poorly differentiated ADC model (Biopsy-poorly ADC model)^25^. The existing model (Biopsy-poorly ADC model) achieved ROC-AUCs in the range of 0.638 - 0.899 on the two independent ESD test sets (Table 3). The existing model (Biopsy-poorly ADC model) has been trained using purely endoscopic biopsy specimen WSIs^25^. Endoscopic biopsy often yields samples that include muscularis mucosae, except in regions (e.g., gastric body) where the mucosal folds are thick. On the other hand, the mucosa surrounding the lesion is circumferentially incised and the submucosal layer is dissected from the proper muscle layer by ESD procedures^9^. ESD specimens usually consist of mucosa, muscularis mucosae, and submucosa with layered tissue architectures^35^. Therefore, there are histopathological differences between endoscopic biopsy and ESD in terms of tissue and cellular components, which might be a primary cause of lower ROC-AUC values in ESD test sets as compared to biopsy test sets by the existing model (Biopsy-poorly ADC model) (Table 3). The deep learning model (ESD-poorly ADC model) was trained by the transfer learning approach from our existing model (Biopsy-poorly ADC model)^25^. We used the partial fine-tuning approach^36^ to train the model faster, as there are less weights involved to tune. We used only 1,120 ESD WSIs (poorly differentiated ADC: 140 WSIs, differentiated ADC: 290 WSIs, non-neoplastic lesion: 690 WSIs) (Table 1) without manual drawing annotations to indicate cancerous tissue areas by pathologists^24,30,37^. After specifically training on ESD WSIs, the model (ESD-poorly ADC model) significantly improved prediction performance on ESD test sets (Table 2) compared to the existing model (Biopsy-poorly ADC model) (Table 3, 4). Importantly, the model (ESD-poorly ADC model) achieved high ROC-AUC (0.929) in surgical specimen test sets (Table 2, 3) and predicted poorly differentiated ADC cell infiltrating area precisely in the serial surgical sections (Fig. 7), which would be very useful to inspect presence or absence of poorly differentiated ADC in massive numbers of surgical serial sections by pathologists in routine clinical workflow. Moreover, the model (ESD-poorly ADC model) also achieved high ROC-AUC values in endoscopic biopsy test sets as compared to the existing biopsy model (Biopsy-poorly ADC model) (Table 2, 3). Thus, for endoscopic biopsy specimens, both deep learning models (ESD-poorly ADC model and Biopsy-poorly ADC model) can classify poorly differentiated ADC precisely, however, the biopsy model (Biopsy-poorly ADC model) achieved slightly better ROC-AUC values (Table 3), so that it would be better to apply the biopsy model (Biopsy-poorly ADC model) for endoscopic biopsy specimens in the routine workflow (Fig. 8).

One of the limitations of this study is that the deep learning models (both ESD-poorly ADC model and Biopsy-poorly ADC model) false negatively predicted poorly differentiated ADC cells (SRCC cells) in foveolar-type SRCC biopsy and ESD WSIs^3^. In early stage, SRCC cells proliferate predominantly in the proliferative zone (near the mucous neck cells)^38^, which were consistently false negatively predicted as poorly differentiated ADC. To predict foveolar-type SRCC precisely, we need to collect a number of foveolar-type SRCC biopsy and ESD cases for additional training or active learning^39^. Another limitation of this study is that it primarily included specimens from a limited number of hospitals and suppliers in Japan, and, therefore, the model could potentially be biased to such specimens. Further validation on a wide variety of specimens from multiple different origins would be essential to ensure the robustness of the model.

The deep learning model established in the present study offers promising results that indicate it could be beneficial as a screening aid for pathologists prior to observing gastric ESD histopathology on glass slides or WSIs. The combination of the deep learning models (ESD-poorly ADC model and Biopsy-poorly ADC model) can cover to predict gastric poorly differentiated ADC precisely in ESD, endoscopic biopsy, and surgical specimen WSIs. At the same time, the model could be used as a double-check tool to reduce the risk of missed poorly differentiated ADC cells. The most important advantage of using a fully automated computational tool as a computer-aided diagnosis is that it can systematically handle large amounts of WSIs without potential bias due to the fatigue commonly experienced by pathologists.

## Materials and methods

### Clinical cases and pathological records

This is a retrospective study. A total of 5,103 H&E (hematoxylin & eosin) stained gastric histopathological specimens (2,506 ESD, 1,866 endoscopic biopsy, and 731 surgical specimen) of human poorly differentiated ADC, differentiated ADC, and non-neoplastic lesions were collected from the surgical pathology files of six hospitals: Sapporo-Kosei General Hospital (Hokkaido, Japan), Kamachi Group Hospitals (Wajiro, Shinyukuhashi, Shinkuki, and Shintakeo Hospitals) (Fukuoka, Japan), and International University of Health and Welfare, Mita Hospital (Tokyo, Japan) after histopathological review of those specimens by surgical pathologists. The cases were selected randomly so as to reflect a real clinical setting as much as possible.

Each WSI diagnosis was observed by at least two pathologists, with the final checking and verification performed by a senior pathologist. All WSIs were scanned at a magnification of x20 using the same Leica Aperio AT2 Digital Whole Slide Scanner (Leica Biosystems, Tokyo, Japan) and were saved as SVS file format with JPEG2000 compression.

### Dataset

Hospitals which provided histopathological specimen slides in the present study were anonymised (Hospital-A-F). Table 1 breaks down the distribution of training and validation sets of gastric ESD WSIs from Hospital-A. Validation sets were selected randomly from the training sets (Table 1). The test sets consisted of ESD, biopsy, and surgical specimen WSIs (Table 2). The patients’ pathological records were used to extract the WSIs’ pathological diagnoses and to assign WSI labels. Training set WSIs were not annotated, and the training algorithm only used the WSI diagnosis labels, meaning that the only information available for the training was whether the WSI contained gastric poorly differentiated ADC or was non-poorly differentiated ADC (differentiated ADC and non-neoplastic lesion), but no information about the location of the cancerous tissue lesions. We have confirmed that surgical pathologists were able to diagnose test sets in Table 2 from visual inspection of the H&E stained slide WSIs alone.

### Deep learning models

We trained the models via transfer learning using the partial fine-tuning approach^36^. This is an efficient fine-tuning approach that consists of using the weights of an existing pre-trained model and only fine-tuning the affine parameters of the batch normalization layers and the final classification layer. For the model architecture, we used EfficientNetB1^40^ starting with pre-trained weights on ImageNet. Figure 1 shows an overview of the training method. The training methodology that we used in the present study was the same as reported in our previous studies^27,29,31^. For completeness we repeat the methodology here. We performed slide tiling by extracting square tiles from tissue regions of the WSIs. We started by detecting the tissue regions in order to eliminate most of the white background. We did this by performing a thresholding on a grayscale version of the WSIs using Otsu’s method^41^. During prediction, we performed the tiling of the tissue regions in a sliding window fashion, using a fixed-size stride (224×224 pixels). During training, we initially performed random balanced sampling of tiles extracted from the tissue regions, where we tried to maintain an equal balance of each label in the training batch. To do so, we placed the WSIs in a shuffled queue such that we looped over the labels in succession (i.e., we alternated between picking a WSI with a positive label and a negative label). Once a WSI was selected, we randomly sampled 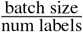 tiles from each WSI to form a balanced batch.

To maintain the balance on the WSI, we oversampled from the WSIs to ensure the model trained on tiles from all of the WSIs in each epoch. We then switched to hard mining of tiles. To perform the hard mining, we alternated between training and inference. During inference, the CNN was applied in a sliding window fashion on all of the tissue regions in the WSI, and we then selected the *k* tiles with the highest probability for being positive. This step effectively selects the tiles that are most likely to be false positives when the WSI is negative. The selected tiles were placed in a training subset, and once that subset contained *N* tiles, the training was run. We used *k* = 8, *N* = 224, and a batch size of 32.

To obtain a single prediction for the WSIs from the tile predictions, we took the maximum probability from all of the tiles. We used the Adam optimizer^42^, with the binary cross-entropy as the loss function, with the following parameters: *beta*_1_ = 0.9, *beta*_2_ = 0.999, a batch size of 32, and a learning rate of 0.001 when fine-tuning. We used early stopping by tracking the performance of the model on a validation set, and training was stopped automatically when there was no further improvement on the validation loss for 10 epochs. We chose the model with the lowest validation loss as the final model.

### Software and statistical analysis

The deep learning models were implemented and trained using TensorFlow^43^. AUCs were calculated in python using the scikit-learn package^44^ and plotted using matplotlib^45^. The 95% CIs of the AUCs were estimated using the bootstrap method^46^ with 1000 iterations.

The true positive rate (TPR) (also called sensitivity) was computed as

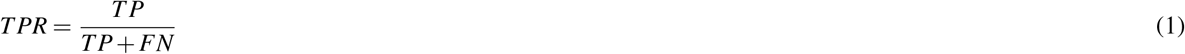

the false positive rate (FPR) was computed as

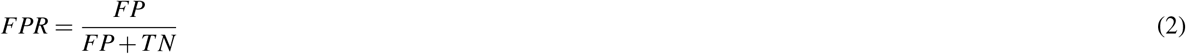

the true negative rate (TNR) (also called specificity) was computed as

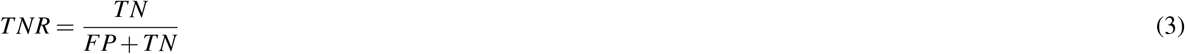

and the accuracy was computed as

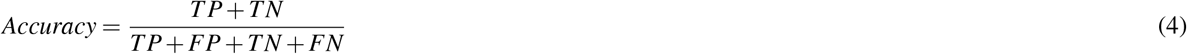

Where TP, FP, TN, and FN represent true positive, false positive, true negative, and false negative, respectively. The ROC curve was computed by varying the probability threshold from 0.0 to 1.0 and computing both the TPR and FPR at the given threshold.

### Availability of data and material

The datasets generated during and/or analysed during the current study are not publicly available due to specific institutional requirements governing privacy protection but are available from the corresponding author on reasonable request. The datasets that support the findings of this study are available from Sapporo-Kosei General Hospital (Hokkaido, Japan), Kamachi Group Hospitals (Fukuoka, Japan), and International University of Health and Welfare, Mita Hospital (Tokyo, Japan), but restrictions apply to the availability of these data, which were used under a data use agreement which was made according to the Ethical Guidelines for Medical and Health Research Involving Human Subjects as set by the Japanese Ministry of Health, Labour and Welfare (Tokyo, Japan), and so are not publicly available. However, the data are available from the authors upon reasonable request for private viewing and with permission from the corresponding medical institutions within the terms of the data use agreement and if compliant with the ethical and legal requirements as stipulated by the Japanese Ministry of Health, Labour and Welfare.

### Code availability

To train the classification model in this study we used the publicly available TensorFlow training script available at https://github.com/tensorflow/models/tree/master/official/vision/image_classification.

## Acknowledgements

This study is based on results obtained from a project, JPNP14012, subsidized by the New Energy and Industrial Technology Development Organization (NEDO). We are grateful for the support provided by Professor Takayuki Shiomi at Department of Pathology, Faculty of Medicine, International University of Health and Welfare; Dr. Ryosuke Matsuoka at Diagnostic Pathology Center, International University of Health and Welfare, Mita Hospital; and Dr. Shigeo Nakano at Kamachi Group Hospitals (Fukuoka, Japan). We thank pathologists who have been engaged in reviewing cases for this study.

## Compliance with Ethical Standards

The experimental protocol was approved by the ethical board of the Sapporo-Kosei General Hospital (No. 576), International University of Health and Welfare (No. 19-Im-007), and Kamachi Group Hospitals (No. 173). All research activities complied with all relevant ethical regulations and were performed in accordance with relevant guidelines and regulations in the all hospitals mentioned above. Informed consent to use histopathological samples and pathological diagnostic reports for research purposes had previously been obtained from all patients prior to the surgical procedures at all hospitals, and the opportunity for refusal to participate in research had been guaranteed by an opt-out manner.

## Contributions

M.T., S.I., and F.K. designed the studies; M.T. and F.K. performed experiments and analyzed the data; M.T. and F.K. performed computational studies; S.I. performed pathological diagnoses, reviewed cases, and pathological discussion; M.T. wrote the manuscript; M.T. supervised the project. All authors reviewed and approved the final manuscript.

